# PREVALENCE AND PREDICTORS OF STRESS AMONG COVID-19 HEALTH WORKERS IN KABWE DISTRICT- A CROSS SECTIONAL STUDY

**DOI:** 10.1101/2021.12.20.21267832

**Authors:** L. L Mukwasa, R. Mutemwa, M. Zambwe, E. Nkhama, P.J Chipimo

## Abstract

**OBJECTIVE:** This study aimed at determining the magnitude of stress among COVID-19 health workers in Kabwe district.

**METHODS:** The study was a cross-sectional study which recruited 138 health care workers managing COVID-19 cases in Kabwe. Data were collected through structured questionnaires and in-depth interviews. Quantitative data was analyzed using SPSS version 16 while qualitative data was analyzed using Nvivo8.

**RESULTS:** The study obtained 100% responses from the respondents and the prevalence of stress among the respondents was 73%. The nurses were more perceived to experience stress compared to the pharmacy personnel (28% vs. 3%). Similarly, women displayed a higher likelihood of experiencing stress compared to men. Lack of support, increased workload and fear were among the factors leading to stress.

**CONCLUSION:** The study went out to determine stress among healthcare workers in Kabwe district. It was established that nurses were more vulnerable than groups. And women were found to be more stressed than men. It is therefore recommended that effective and meaningful interventions be put in place to mitigate the impact of long-term psychological distress and physical well-being in healthcare workers during the COVID-19 pandemic and future outbreaks.

**ARTICLE SUMMARY:** *Strengths and limitations of the study:*

- A cross-sectional study method was employed; it does not assist in determining the cause and effect, in addition, the timing of the snapshot may not guarantee representation of the situation overtime. Therefore, there may be need to evaluate the effects of the COVID-19 pandemic on health workers mental requirements using a longitudinal study design.
- There might have been bias from respondents as the outcomes were self-reported. However, irrespective of the aforementioned limitations, this is a novel study in Kabwe describing stress among health workers in the district during the COVID-19 pandemic.
- The study grants access to initial evidence on stress among health workers managing the COVID-19 pandemic in Kabwe district, with the expectation of drawing the attention to legislators, health facility supervisors and those involved in the response to COVID-19 or impending epidemics.

## INTRODUCTION

Severe Acute Respiratory Syndrome Coronavirus-2 (SARS-CoV-2) the etiological agent for coronavirus disease 2019 (COVID-19) was first reported from China on 31^st^ December, 2019 as a cause of pneumonia.^1^ It subsequently spread worldwide in less than three months, with Africa reporting its first case, on the 14^th^ of February 2020, in Egypt. Since then, the virus had been detected in all the African countries. This led to the World Health Organization (WHO) to declare the disease, a worldwide pandemic, on the 11^th^ of March 2020.^2^ COVID-19 is generally a severe fatal disease that may result in death owing to progressive respiratory complications.^3, 4^ A suspected case of COVID-19 was defined by the Zambia National Public Health Institute (ZNPHI) in four ways: 1) an acute respiratory illness in a person with a history of international travel in the 14 days prior to symptom onset; 2) an acute respiratory illness in a person with a history of contact with a person with laboratory-confirmed COVID-19 in the 14 days prior to symptom onset; or 3) severe acute respiratory illness requiring hospitalization; or 4) being a household or close contact of a patient with laboratory-confirmed COVID-19.^5^

Health Care Workers (HCWs) have been reported to be exposed to infections on a daily basis, which could result in a significant level of mental stress at work. This is especially true when a global outbreak of an infectious disease occurs. During the Severe Acute Respiratory Syndrome (SARS) outbreak, studies on health care workers indicated severe emotional distress in 18%–57% of workers, which was linked to fear of contagion, concern for family, job stress, and attachment insecurity.^6^ Healthcare workers were among those infected during the SARS outbreak, with some succumbing to the disease, however, the situation has proven to be worse during the COVID-19 pandemic, potentially putting health workers under a lot of stress.^7^

The first cases of COVID-19 in Zambia were recorded on the 18^th^ of March, 2020, in Lusaka^5^ and later spread countrywide. Zambia, like the rest of the world, quickly implemented Public health measures to prevent the spread of the disease. Measures included; frequent hand-hygiene, social distancing, wearing of face masks, closure of public places, heightened disease surveillance at all ports of entry, strengthening of the emergency preparedness and response systems, which included activation of the Public Health Emergency Operations Centre (PHEOC)^8,9^.

Kabwe, a district in Central Zambia, reported its first COVID-19 case on May 18, 2020, and the number of cases has continued to rise since then.^10^ A significant spike in severe COVID-19 cases and deaths was reported in December 2020 and early January 2021, and the second wave of the disease was confirmed on December 30, 2020.^11^This was largely, attributed to the huge gatherings during the festive season, poor adherence to public health guidelines on COVID-19 and a new strain the country was undergoing at the time.^12^ Amid low public compliance on COVID-19 preventive measures across the country, to prevent the potential spread of a deadlier strain of the virus as displayed in the second wave, the Zambia Ministry of Health was compelled to expand testing capacity, disease surveillance and mitigation techniques.^12^ As a result of this, HCWs working for the Ministry of Health in Kabwe, particularly those directly handling the epidemic in the district, got agitated. This meant that they were expected to handle unexpectedly high workloads, longer working hours, insufficient personal protective equipment (PPE), a lack of specific drugs and methods to treat COVID-19, and being separated from family during quarantine, all of which put them at risk for stress-related disorders.^13^ However, to the best knowledge of the authors, there was no prevailing literature that determined the prevalence of stress or its predictors among health workers managing COVID-19 cases in Kabwe district. Therefore, the prevalence of stress and its predictors for these health care workers remained unknown.

This paper reports on the prevalence and predictors of stress among HCWs in Kabwe, managing the COVID-19 pandemic. It is expected that the findings will contribute to a better understanding of the psychological impact of the outbreak on HCWs and will be useful in the planning of stress management among health care workers involved in the fight against COVID-19 and future outbreaks.

## METHODS

A cross sectional mixed research method conducted between December 2020 and 31 March, 2021. The primary technique employed during the study was sequential data collection method, with qualitative data collected through literature review and in-depth interviews, while, quantitative data was collected using of structured questionnaires.

The study population consisted of health workers managing Covid-19 in Kabwe district A list of 215 health workers who were involved in the management of COVID-19 in Kabwe was availed to the researcher and from this list 138 were eligible to participate in the study. This included frontline healthcare professionals, supervisors and support staff; from Kabwe District health Office (KDHO), the Kabwe Rapid Response Team (RRT), frontline health workers working in Covid-19 Isolation Wards at Kabwe Central Hospital and Bwacha isolation centre.

The sample size was obtained using the formula adapted from Evans at al. (2000);

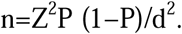

**Where:** n = sample size:

Z = confidence interval (95% = 1.96; Z-score):

P = proportion of interest = estimated at 50%:

d = maximum allowable error ± 5%

**Thus:**

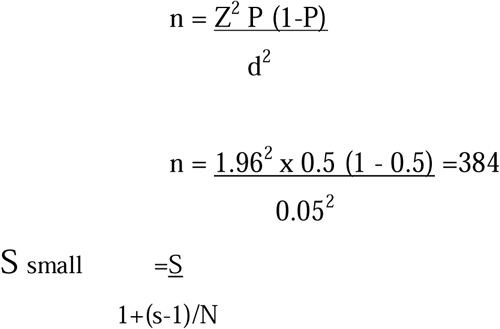

Where: S is the bigger sample size,

s is the small sample size,

N is the total is the total population size.

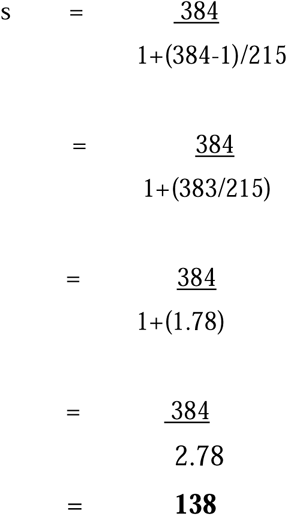

A systematic sampling strategy was adopted in the investigation. The sampling interval was determined by dividing the entire number of health workers managing COVID-19 in Kabwe (215) by the sample size (138). After selecting a number between 1 and the sample interval from the random table, names were assigned numbers; number one being chosen as the beginning point. Then, after skipping one name, two more names were chosen until the appropriate sample size was obtained. Both quantitative and qualitative data were collected from the same sample.

All participants provided informed consent, which stated that the respondent’s anonymity would be maintained and that data would be collected using codes.

### Sampling Criteria

i. **Inclusion criteria**
  a. Frontline health workers working in the district’s Covid-19 rapid response team since 18^th^ May, 2020.
  b. Frontline health workers managing Covid-19 at Kabwe Central Hospital and the Bwacha isolation Centre
  c. Management team and support staff from Kabwe district Health Office, Kabwe Central Hospital
ii. **Exclusion criteria**
  a. Healthcare workers and support staff working in health facilities in Kabwe district but not directly involved in handling Covid-19 cases
  b. Management members not directly involved in coordinating frontline health care workers managing Covid-19 cases

In this study, a dependent variable and independent variables were used. The dependent variable was stress status (yes/no). The independent variables were demographic factors (included, age, sex, marital status, presence of underlying condition) and elements of work characteristics (included department of work, work experience, organisational support, workload, work environment).

Microsoft Excel was used for checking quantitative data completeness and consistencies, Statistical Package for Social Sciences (SPSS) version 16 was used for analysis. Relationships between the dependent and independent variables were assessed using logistic regression. Demographic and work characteristics were summarized using descriptive statistics (frequencies and percentages). Prevalence of stress was expressed with 95% confidence intervals (CI) and statistical significance set at P>0.05. Qualitative data, derived from in-depth interviews was examined using Nvivo8 and content analysis. Responses were grouped and codes created through categorical data. Codes were then reviewed and revised into themes and presented with regards to the second objective of the study. Data collection tools were organized and secured with access restricted only to the research team.

## DEFINITIONS OF OPERATIONAL TERMINOLOGIES

### Prevalence

The number of individuals with the condition (stress) at a specific time

### Stress

a state of mental or emotional strain or tension resulting from adverse or demanding circumstances

### Predictor

something such as an event or fact that enables you to say what will happen in the future

### COVID-19

Severe acute respiratory syndrome coronavirus 2 (SARS-CoV-2) is a novel severe acute respiratory syndrome coronavirus.

### Personal Protective Equipment

Equipment that will protect the user against health or safety risks at work. It can include items such as safety trivet suits, gloves, eye protection, face shields, safety footwear and respiratory protective equipment (RPE).

### Health Workers

One who provides direct care and services to the sick and injured, either as a professional or indirectly as aides, helpers, or medical waste handlers.

## RESULTS

In this study 73% of respondents reported to be stressed, with female respondents accounting for a higher proportion than males (50% vs. 23%). Nurses represented the largest number of the respondents that reported stress (38(28%)) followed by Environmental Health Practitioners (24(17%)). Among the respondents 88(64%) reported to have not received psychological support from their institutions.

**Figure 4:**
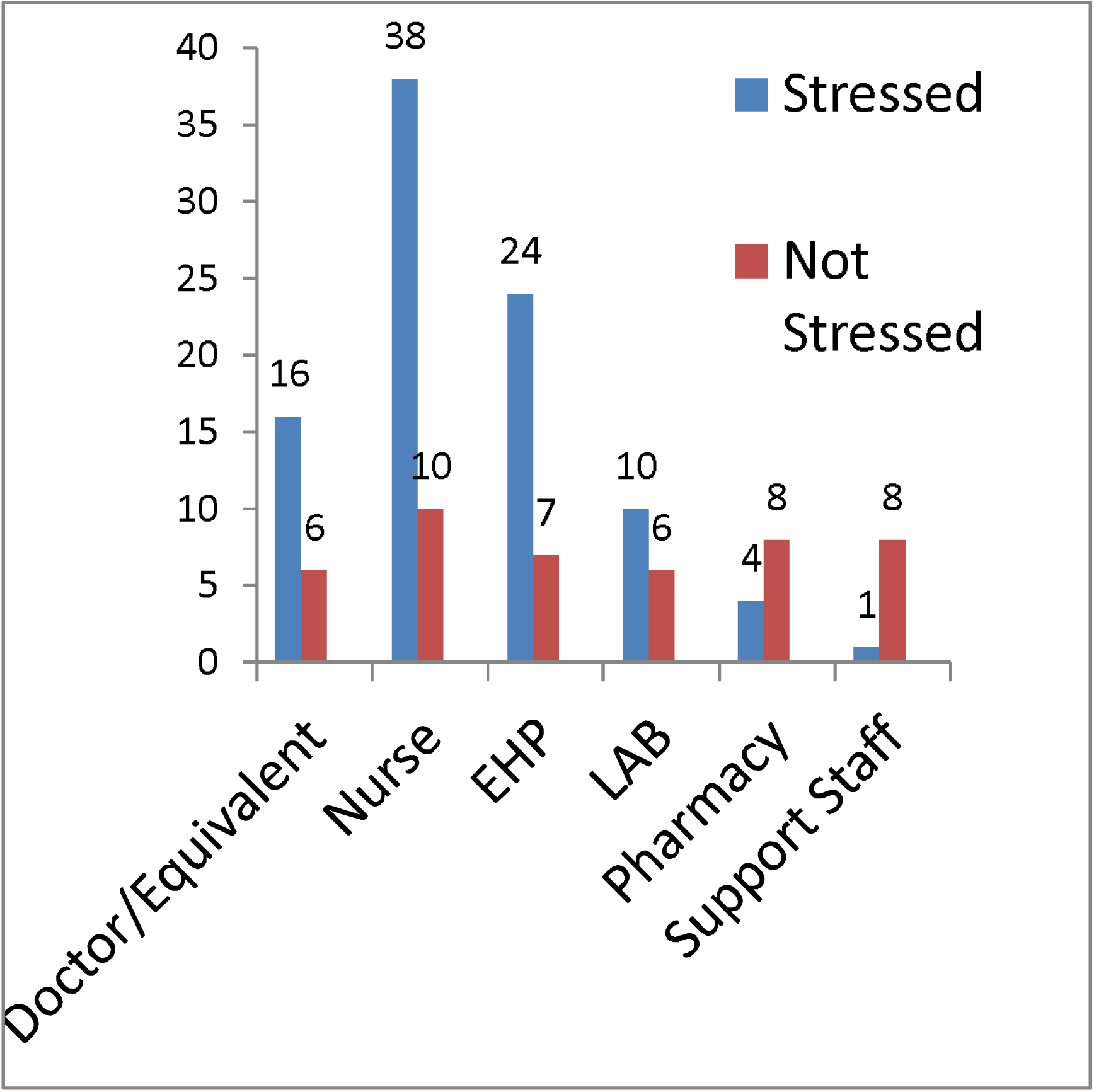
Prevalence of stress according to profession.

**Table 1:** Association between hypothesized predictors and Stress among Health Workers.

| Characteristic | Category | Stress |  | Odds Ratio | 95% CI | P-value (<0.05) |
| --- | --- | --- | --- | --- | --- | --- |
|  |  | Yes | No |  |  |  |
| Sex | Female | 31 | 24 | (Ref) | (Ref) | (Ref) |
|  | Male | 69 | 14 | 7.5704 | (3.24-7.64) | **0.0001** |
| Age | <35 | 53 | 27 | (Ref) | (Ref) | (Ref) |
|  | >=35 | 47 | 11 | 0.4594 | (0.5470 – 2.37) | **0.0578** |
| Co-morbidity | With Co-morbidity | 13 | 5 | (Ref) | (Ref) | (Ref) |
|  | Without Co-morbidity | 82 | 31 | 1.0463 | (0.35 - 3.17) | **0.9361** |
|  | Unknown | 5 | 2 | 1.0400 | (0.15 - 7.22) | 0.9683 |
| Profession | Doctor/Equivalent | 16 | 6 | (Ref) | (Ref) | (Ref) |
|  | Nurse | 38 | 10 | 19.891 | (2.40 -164.66) | **0.01** |
|  | EHOs/EHTs | 24 | 7 | 9 | (3.57- 38.67) | 0.010 |
|  | Lab personnel | 10 | 6 | 11.751 | (0.66 - 11.62) | 0.017 |
|  | Pharmacy | 4 | 8 | 2.78 | (0.05 - 1.36) | 0.010 |
|  | Support staff | 8 | 1 | 0.25 | (0 – 0.3) | 0.018 |
|  |  |  |  | 0.02 |  |  |
| Work experience | Less than 5years | 24 | 9 | (Ref) | (Ref) | (Ref) |
|  | 5-10 years | 40 | 19 | 0.6349 | (0.68 – 4.68) | **0.2831** |
|  | Above 10 years | 36 | 10 | 0.257 | (0.44 – 1.20) | **0.216** |
| Facility Preparedness | Facility is prepared | 16 | 7 | (Ref) | (Ref) | (Ref) |
|  | Facility is not prepared | 84 | 31 | 0.8435 | (0.32 - 2.25) | **0.7334** |
| Psychological Support | Offered Support | 12 | 5 | (Ref) | (Ref) | (Ref) |
|  | Not offered support | 88 | 33 | 0.0207 | (0.01 – 0.06) | **0.0001** |

**Table 2.** Response from the In-depth Interview.

| Sub-themes | Codes | Quotations |
| --- | --- | --- |
| Workload | Heavy workload<br>Unpredictable work schedules | *I was ever tired and very fatigued but was not allowed to rest due to the unpredictability of work schedule and shortage in Human resource (EHT). We were expected to screen patients, dispense drugs and at the same time offer nursing care (Nurse).* |
| Losing control over situation | Losing control<br>*Lack of specific treatments*<br>*Reduction of existing human resources* | *As cases increased during the second wave we started feeling a lot like were losing control due the reason that the disease had no cure and affected individuals differently. There was no standard treatment, Human resource was inadequate and some had very limited work experience, it was more of a job on training for them(Nurse)* |
| Inadequate protective equipment and the difficulty in using them | *Shortage in personal protective equipment and difficulty of usage for long periods* | *We never really had adequate PPE, In the same suit I would test over 200 people in one day in different settings without proper disinfection of the suit. It was also uncomfortable to wear for a long time due to the nature of the material used for the suits, I would sweat very much. (Lab Personnel)*<br>*N95 masks and protective wear is only reserved for Lab personnel during contact tracing, we were only given surgical masks to protect ourselves.(EHT)* |
| Uncertainty of diagnosis | Inability to distinguish | *Minus the test being done one is unable to distinguish between Covid and non Covid patients, due to the similarities in the presented* |
|  | between Non-Covid and Covid patients | *symptoms with other diseases and the turnaround time for the test results is very long. This often leaves us feeling anxious and helpless (Physician)* |
| Sense of powerlessness | Managing cases with no definite treatment | *I often feel anxious about exposing my children, other family members and friends and the possibility of infecting them leaves me with a sense of powerlessness it seems you can just never be too careful (Nurse)* |
| A sense of care and self-sacrifice | *Self-sacrifice and Loyalty to the medical oath* | *We want to save anyone who gets sick despite the fact that we place ourselves in harm's way. We took the oath and cannot go back on it, although sometimes we feel like giving up, we have to wake up and soldier on (physician).* |
| Ambiguity | *Disapproval of new drugs* | *Since Covid-19 is a new disease and no one knows exactly what to do. A lot of drugs have been introduced whilst some dispute their effectiveness others rely on them and this is very confusing. One day you are told to use a certain drug and the next day you are told to stop, it all a trial and very frustrating (Pharmacist)* |

## DISCUSSION

The majority of participants (83 (66.1%)) were females between the ages of 25 and 34, were married and a large proportion reported having experienced stress. The high number of female participants experiencing stress could partly be explained by the fact that they had families whom they were concerned about infecting with COVID-19, causing stress, which was widely reported among this group of respondents. The female respondents had a 7-fold higher risk of developing stress than their male counterparts.These findings are similar to a study conducted in China, were, an analysis of the risk factors for stress, depression, and anxiety symptoms revealed that female gender was a significant predictor, with women having a 2-fold increased risk of developing these conditions.^14^ Age and Marital status had no relationship with stress in this study, neither did the indicator of having an underlying condition (co-morbidity) affect the stress status of the respondent (P>0.08).

According to this study, one of the significant relationships of the hypothesized predictors of stress among the health workers was profession (P>0.0001), with health professionals having a higher risk of developing stress. Nurses had the highest proportion of respondents (28%) who reported being stressed and had a higher likelihood of experiencing stress (OR 19). This could be attributed to the heavy workload that Kabwe nurses face. They are expected to not only spend more time in patient care by screening and dispensing drugs, but also to provide full-time nursing care. This finding is consistent with the findings of a study conducted in Nepal on the mental health impacts of health workers during COVID-19 in a low-resource setting.^15^ Similar findings were reported in another study conducted in China, primarily attributed higher likelihood to nurses spending more time in patient care than other health workers.^14^

It is a well-known fact that providing adequate PPE to HCWs boosts their confidence and resilience to psychological problems, which has an effect on healthcare service delivery during the COVID-19 pandemic. One of the causes of psychological anguish among the 110/138 respondents in this study who felt they were not adequately protected and were vulnerable to COVID-19 infections was inadequate provision of PPE by health authorities. This is consistent with findings from other studies, which emphasized the need of providing health workers with proper PPE as well as psychological support in order to promote resilience to negative mental health outcomes.^16,17^ Work experience and work environment were not significantly associated with stress in this study (P>0.3 and P>0.7, respectively). This indicates that, regardless of the number of years the respondents worked or the alignment of their work environment, feeling stressed was unrelated to either event. This contradicts the findings of a study conducted in Ethiopia, were work-related stress was related to working experience.^18^ However, these findings are consistent with a study conducted in Saudi Arabia, were work experience had no effect on workers’ stress levels.^19^ The fact that the work environment, such as facility preparedness, may not cause stress in workers may imply that other factors, other than the work environment, may be at work in contributing to stress levels in the respondents.

In this study, 73% of health workers reported feeling stressed. All of the respondents were concerned about the possibility of spreading the disease to their loved ones; some had young children under the age of five, and others lived with elderly relatives, friends, or family members who had underlying conditions that put them at high risk of severe infections. This is consistent with the findings of similar studies in Wuhan, China, assessing the psychological impact of the COVID-19 pandemic, which established that stress was prevalent in 29.8% of healthcare professional.^20^ It is also consistent with another study, conducted in New York (United States of America), which indicated that healthcare workers treating patients with COVID-19 at the epicenter, experienced distress, ranging from fears of COVID-19 transmission, to concerns about family and home life.^21^

Concerns about a lack of institutional psychological support (88%) were also expressed and cited as demotivating factors by respondents. This study’s findings indicate that a lack of psychological support from the institution contributed to the high prevalence of stress among health workers (P>0.001) and is consistent with the findings of another study, which found that “greater social support was associated with lower perceived stress during the early phase of the COVID19 pandemic.^22^

Respondents reported having less or no time for family and friends as a result of working extremely long hours that frequently left them exhausted, as reported in previous studies^23^. To cope with stress, the majority of respondents relied on spending time with or talking to family and friends. The majority of those interviewed expressed anxiety about the COVID-19 pandemic’s unclear future. They feared that if the situation got out of hand, the country’s health-care system would not be able to handle it, leading to despair and the fear of the disease not being eradicated.

## CONCLUSION

This study was successful in determining the prevalence of stress among health workers managing COVID-19 in Kabwe district and identifying its predictors. It was revealed that approximately 73% of the health workers reported being stressed by the COVID-19 situation. This is not surprising given that health workers are also human, and the pressure from health authorities and the general public to provide quality healthcare services in the midst of a pandemic may make them vulnerable to stress. Sex, organizational psychological support, and profession were found to be statistically significant predictors of stress. Stress was also exacerbated by insufficient PPE, a heavy workload, unpredictable work schedules, and uncertainty about the COVID-19 situation. This implies that healthcare workers should be given special consideration due to their high risk of developing psychological problems. The study’s findings are expected to inform the development and implementation of interventions to reduce the effects of continuous stress on healthcare workers’ well-being. Furthermore, the lessons learned from the COVID-19 pandemic are hoped to help decision-makers at all levels of government, hospital management, and the community influence emergency preparedness policies and promote readiness to protect healthcare workers not only from Covid-19, but also from future public health crises.

### 6.1 RECOMMENDATIONS

- Health care institutions, such as district health offices and hospitals, must develop appropriate strategies to investigate stress in health care settings.
- It is critical to develop interventional programs to identify and alleviate stress sources and effects, such as routine screening for psychological problems and early intervention.
- Health institutions at the district and hospital levels should create an enabling work environment with a good support system, adequate availability of PPE, proper training of health workers on COVID-19 management, and a focus on incentives that boost their work morale.
- Staff and management must be given time and space for reflection and support in order to appreciate the need of maintaining their own resilience in the face of a public health catastrophe.
- In order to provide support to members of staff and enhance working conditions, staff training on current public health concerns is required.
- Using a longitudinal study method, more research is needed to identify the causal factors that contribute to the prevalence of stress among healthcare personnel

## Supporting information

strobe check list

## Data Availability

All data produced in the present study are available upon reasonable request to the authors

## ETHICS APPROVAL AND DISSEMINATION

Approval to conduct the study was obtained from the University of Lusaka, School of Medicine and Health Sciences Research Ethics Committee; Reference: IORG0010092/MPH18212932. Thereafter, authorization was obtained from the Ministry of Health-Kabwe District Health Office and Kabwe Central Hospital. The end study results will be published in an open access peer-reviewed journal.

## DATA SET ACCESS

The Data set not available online. Nonetheless, it may be made available to third parties upon reasonable request to the corresponding author.

## AUTHOR CONTRIBUTIONS

LM

- Work conception. Data acquisition, analysis and interpretation.
- Manuscript drafting.
- Approval of final manuscript.
- Accountable for all aspects of the work regarding its accuracy or integrity.

RM

- Work conception, analysis and interpretation.
- Manuscript drafting
- Approval of final manuscript.
- Accountable for all aspects of the work regarding its accuracy or integrity

MZ

- Data interpretation
- Manuscript drafting and critical revisions for intellectual content.
- Approval of final manuscript.
- Accountable for all aspects of the work regarding its accuracy or integrity

EN

- Data interpretation
- Manuscript drafting and critical revisions for intellectual content.
- Approval of final manuscript.
- Accountable for all aspects of the work regarding its accuracy or integrity

PJC

- Data interpretation
- Manuscript drafting and critical revisions for intellectual content.
- Approval of final manuscript.
- Accountable for all aspects of the work regarding its accuracy or integrity

## ACKNOWLEDGEMENTS

This article is a part of the Master’s thesis submitted to the University of Lusaka in partial fulfillment of the requirements for the degree of Master of Public Health

The District Health Director for Kabwe, Dr. Jerry Sinyangwe, and the Medical Superintendent for Kabwe Central Hospital, Dr. Victor Kusweji, deserve our sincere gratitude and appreciation for allowing this study to take place. Special thanks go to all of the health care professionals who participated in this study.

Finally, we would like to thank everyone who contributed to this study but was not specifically mentioned.

